# Impact of patient engagement on the design of a mobile health technology for cardiac surgery

**DOI:** 10.1101/2020.07.22.20159848

**Authors:** Anna M. Chudyk, Sandra Ragheb, David E. Kent, Todd A. Duhamel, Carole Hyra, Mudra G. Dave, Rakesh C. Arora, Annette S.H. Schultz

## Abstract

**Objective:** The aims of this study were to describe the impact of patient engagement on the initial design and content of a mobile health (mHealth) technology that supports enhanced recovery protocols (ERPs) for cardiac surgery.

**Methods:** Engagement occurred at the level of consultation and took the form of an advisory panel. Patients that underwent cardiac surgery (2017-2018) at St. Boniface Hospital (Winnipeg, Manitoba) and consented to be contacted about future research, and their caregivers, were approached for participation. A qualitative exploration was undertaken to determine advisory panel members’ key messages about, and the impact of, patient engagement on mHealth technology design and content.

**Results:** Ten individuals participated in the advisory panel. Key design-specific messages centered around access, tracking, synchronization, and reminders. Key content-specific messages centered around roles of cardiac surgery team members and medical terms, educational videos, information regarding cardiac surgery procedures, travel before/after surgery, nutrition (i.e., what to eat), medications (i.e., drug interactions), resources (i.e., medical devices), and physical activity (i.e., addressing fears and providing information, recommendations, and instructions). These key messages were a rich source of information for mHealth technology developers and were incorporated as supported by the existing capabilities of the underlying technology platform.

**Conclusions:** Patient engagement facilitated the development of a mHealth technology whose design and content were driven by the lived experiences of cardiac surgery patients and caregivers. The result was a detail-oriented and patient-centered mHealth technology that helps to empower and inform patients and their caregivers about the patient journey across the perioperative period of cardiac surgery.

**KEY QUESTIONS:** 

**What is already known about this subject?:** Enhanced recovery protocols (ERPs) have been proposed as a clinical strategy to effectively address complex and multi-system vulnerabilities, like those commonly present in older adults undergoing cardiac surgery. Mobile health (mHealth) technologies have the potential to improve delivery and patient experience with ERPs, but their development in the academic research setting is often limited by a lack of end-user (e.g., i.e., patient and caregiver) involvement.

**What does this study add?:** To our knowledge, this is one of the first studies to engage patients and caregivers in the development of a mHealth technology that supports ERPs for cardiac surgery. This study describes a process for engaging patients and caregivers as “co-producers” of a mHealth technology to support delivery of ERPs during the perioperative period of cardiac surgery. It also demonstrates that engaging patients and caregivers in research, through the formation of an advisory panel, yields a rich source of information to guide the design and content of mHealth technologies in cardiac research.

**How might this impact on clinical practice?:** In an era in which mHealth technologies are being increasingly looked to for the optimization of healthcare delivery, this study underscores the utility of using patient and caregiver voices to drive the development of patient-centered mHealth technologies to support clinical practice.

## INTRODUCTION

Enhanced recovery protocols (ERPs) are evidence-based care pathways aimed at standardizing perioperative care. In offering a multimodal and interdisciplinary approach to perioperative care, they have been proposed as a clinical strategy to effectively address complex and multi-system vulnerabilities,[1, 2] like those commonly present in older adults undergoing cardiac surgery.[3, 4] Mobile health (mHealth) refers to medical and public health practice supported by mobile devices (e.g., smart phones, tablets, patient monitoring devices, etc.).[5] Mobile health technologies, such as application-based platforms (a.k.a. “apps”), have the potential to enhance the utility of ERPs through increasing the effectiveness of information delivery and patients’ (and caregivers’) retention of information regarding their health care plan.[6, 7] There is some evidence to support the feasibility of using mHealth technology with cardiac surgery patients during their inpatient recovery.[8] However, researchers’ efforts to design mHealth technologies for the perioperative cardiac surgery setting (and in general) are commonly limited by their failure to involve end-users (such as patients and caregivers) in research activities.[9]

Patients and caregivers may be involved in mHealth technology design studies as research participants or collaborators. This latter approach, which involves the “co-production” of research with patients and caregivers, is a form of participatory research commonly referred to as patient engagement, patient and public involvement, patient involvement, or stakeholder engagement in research. In this study, we use the term patient engagement in research, and define it as the formation of meaningful and active collaborations between researchers and patients (and informal caregivers) in research governance, priority setting, conduct, and/or knowledge translation.[10] The importance of co-producing research with patients and caregivers can be viewed both from a moral perspective (e.g., those affected by an issue should be actively involved in the generation of solutions to it),[11, 12] as well as in terms of its potential to improve the methodological quality, relevance, and/or uptake of research.[13, 14, 15] Moreover, lack of attention to users’ perspectives during the design phase is one of the competing explanations for the relatively low uptake of mHealth technologies by patients.[9] An important step towards a more wide spread adoption of patients and caregivers as co-producers of mHealth technology research is one that facilitates a better understanding of processes for engaging patients and caregivers in mHealth technology design in the research setting.

This study was set within the context of a Canadian clinical research hospital where our research group is involved in the development and implementation of ERPs for cardiac surgery. One component of this larger project aims to adapt a mHealth technology and determine its effectiveness in improving knowledge delivery and patient adherence with ERPs during the perioperative period of cardiac surgery. In this manuscript, we focus on the patient engagement process employed to adapt the mHealth technology, which was guided by the Canadian Institutes of Health Research’s (CIHR’s) Patient Engagement Framework [10] and our scoping review of models and frameworks of patient engagement in health services research.[16] Given the novelty of engaging patients as co-producers of mHealth technology in the academic research setting and among most of our team members, this manuscript’s aims were to describe the process and impact of patient engagement on the initial design and content of a mHealth technology that supports ERPs for cardiac surgery.

## METHODS

This study was set in an academic, tertiary care centre that performs cardiac surgery (St. Boniface Hospital, Winnipeg, Manitoba). Ethics approval for the study was obtained from the University of Manitoba Research Ethics Board as well as the Research Review Committee at St. Boniface Hospital. Patients and caregivers provided written consent and were compensated $50 (for time and transportation) in addition to the cost of parking per meeting that they attended. The Guidance for Reporting Involvement of Patients and the Public (GRIPP2) long-form checklist guided the reporting of patient engagement in this article.[17]

### Overview of mHealth technology

The mHealth technology under development was an app-based platform hosted by BeeWell Health ©.[18] For this study, the intention was to adapt and electronically format patient derived content that addressed the cardiac surgery patient journey from initial surgery consent through to the eight-week post-operative recovery period, for delivery via the mHealth technology. This content sought to target three aspects of perioperative care (i.e., patient tailored education, optimization of patient health, and patient engagement in care) and was focused on four domains of information (i.e., nutrition, medications, resources and physical activity). A screenshot from the mHealth technology is provided in Figure 1.

**Figure 1.**
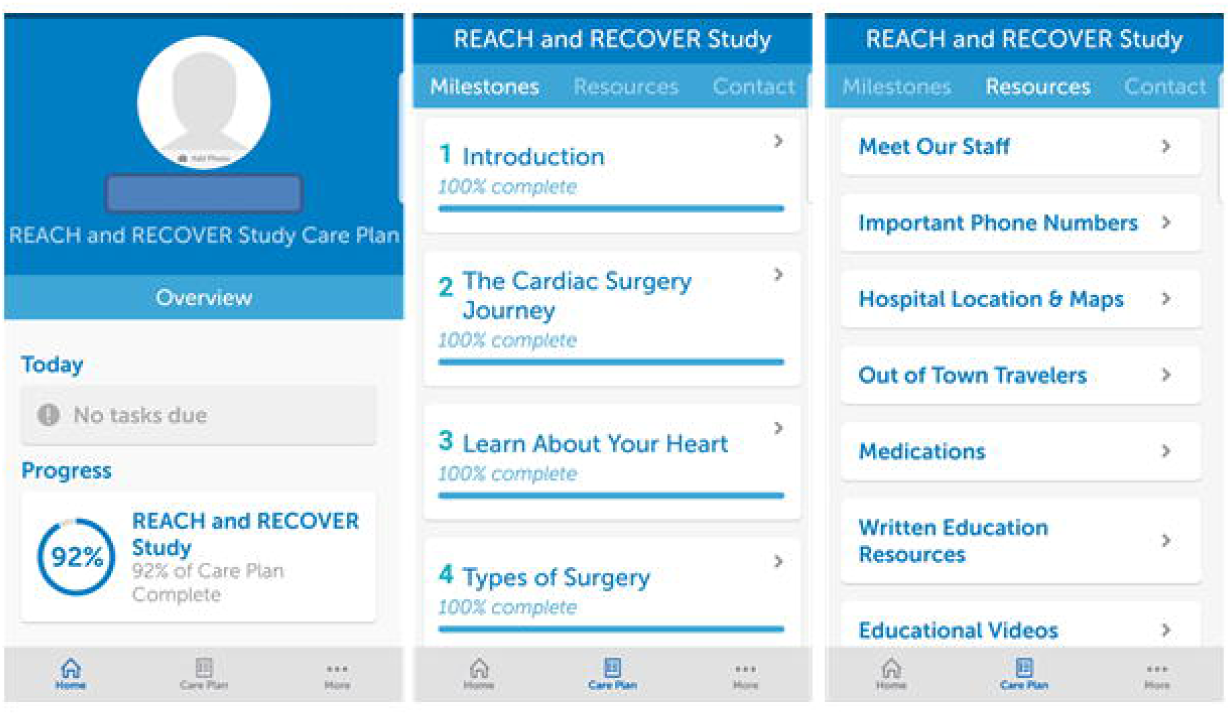
Screenshot from the mobile health technology.

### Patient involvement

The four domains of information targeted by our adaptation of the mHealth technology were informed by our previous work (data unpublished) with cardiac surgery patients and caregivers. Specifically, focus group sessions identified these areas as cardiac surgery patient and caregiver priorities. Continued research (i.e., online and telephone surveys) validated these findings within a larger patient and caregiver population. Patients and caregivers were involved as co-researchers in the conduct of this study, as expanded upon below, and were all invited to co-author this manuscript.

### Description of patient engagement process

Patient engagement in research encompasses a wide range of activities and participation types, as influenced by the characteristics of a given project (e.g., scope, time, and financial resources) and the contributions patients are willing to offer (see Supplementary Material 1 for an overview of patient engagement in research).[10, 13, 14, 19] In the present study, engagement took the form of an advisory panel and occurred at the level of consultation.[19] The role of the advisory panel was to inform the design and content of the mHealth technology. The advisory panel met in-person three times, approximately two weeks apart. Each meeting was approximately three hours in duration. Figure 2 displays an outline of activities that occurred at each meeting.

**Figure 2.**
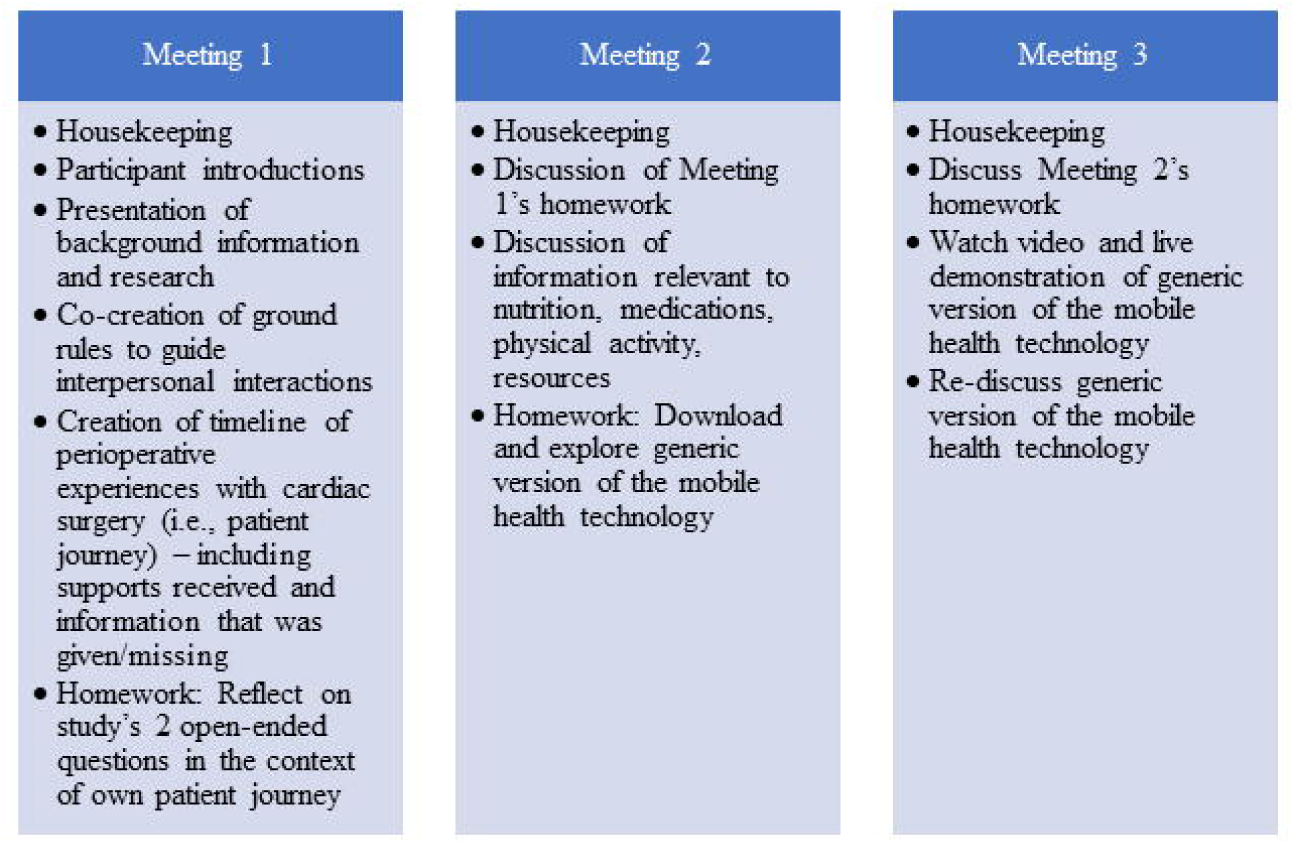
Outline of activities by meeting.

The activities that occurred within the meetings were not only developed to gather advisory panel input on the design and content of the mHealth technology, but also to create/facilitate an environment that supported the guiding principles that underlie patient engagement (i.e., mutual respect, inclusiveness, co-building, support; see Supplementary Material 2 for information on the employed approach to creating an environment that embodied these guiding principles).[10] The primary method for obtaining panel members’ input on the design and content of the mHealth technology was group discussions. These discussions centered around two open-ended questions --“*what information stuck out as important during your patient journey*” and “*what information do you wish you had known during your patient journey*”. In addition, the scope of the discussions was narrowed to the four domains of information (i.e., nutrition, medications, resources and physical activity) identified through previous work, as well as to the content and layout of information presented in a downloadable generic version of the mHealth technology. A skilled facilitator (DEK) led the meetings based on a developed facilitation guide. A notetaker (MD) and an audio recorder documented meeting proceedings.

### Recruitment

Patients who underwent a cardiac surgery procedure within the previous two years (2017-2018) at the study hospital and consented to be listed in a database of individuals interested in participating in future research, and their caregivers, were approached for advisory panel membership. Panel members were selectively chosen for diversity in sex and procedure type and were excluded if they could not read and/or communicate in English. Recruitment was targeted at 10-12 individuals based on ours and others’ [20] experiences with group dynamics and group size.

### Impact of patient engagement

A qualitative exploration was undertaken to determine the impact of patient engagement on the design and content of the mHealth technology. This included description of (a) the key messages generated by the advisory panel, (b) how key messages were incorporated into the development of the mHealth technology, and (c) feedback from the developers of the mHealth technology about their experience with using the information generated by the advisory panel to guide development of the mHealth technology.

### Data analysis

Discussions that occur as part of patient engagement activities do not typically produce data that are thematically analyzed,[21] as the purpose of patient engagement is to learn from patient experiences, not interpret patient experiences through the researcher’s lens. Thus, “real-time processing” of information takes place during discussions and the information that is gathered is generally presented as a list of patient-made recommendations used to support project decision-making.[21] Accordingly, the meeting facilitator (DEK) employed common techniques (e.g., summarization, reflection, asking clarifying questions) to identify advisory panel members’ key messages during discussions. Two study team members (DEK and AMC) reviewed the research assistant’s notes from all three meetings, along with transcripts from the second meeting, to generate a list of key messages about design and content of the mHealth technology. These key messages were presented by a study team member (DEK) to the developers of the mHealth technology platform to guide the design and content of the mHealth technology.

## RESULTS

Ten individuals (six patients and four caregivers) participated in the advisory panel. Select sociodemographic characteristics of advisory panel members are shown in Table 1. Each caregiver (n=4) was a patient’s (n=4) spouse. Two of the patients did not have a caregiver attend any of the advisory panel sessions. A summary of advisory panel members’ key messages about the design and content of the mHealth technology follows.

**Table 1.** Select sociodemographic characteristics of advisory panel members. All results are presented as number (%) or median (25^th^ percentile, 75^th^ percentile).

| Variable | Patients (n=6) | Caregivers (n=4) |
| --- | --- | --- |
| Age (years) | 74 (72, 76) | N/A<sup>b</sup> |
| Female | 3 (50%) | 3 (75%) |
| **Ethnicity** |  |  |
| White/Caucasian/European | 5 (83%) | 4 (100%) |
| First Nations/Inuit/Metis | 1 (17%) | 0 (0%) |
| **Procedure type** |  |  |
| Aortic valve replacement | 3 (50%) | N/A<sup>b</sup> |
| Aortic valve replacement/CABG<sup>a</sup> | 1 (17%) | N/A<sup>b</sup> |
| Aortic valve replacement/<br>mitral valve replacement | 1 (17%) | N/A<sup>b</sup> |
| mitral valve replacement | 1 (17%) | N/A<sup>b</sup> |

<sup>a</sup> Coronary artery bypass grafting
<sup>b</sup> Not applicable

### Key messages about mHealth technology design

Key messages about the design features of the mHealth technology related to access, tracking, synchronization, and reminders. Specific key messages about mHealth technology design are in Table 2.

**Table 2.**
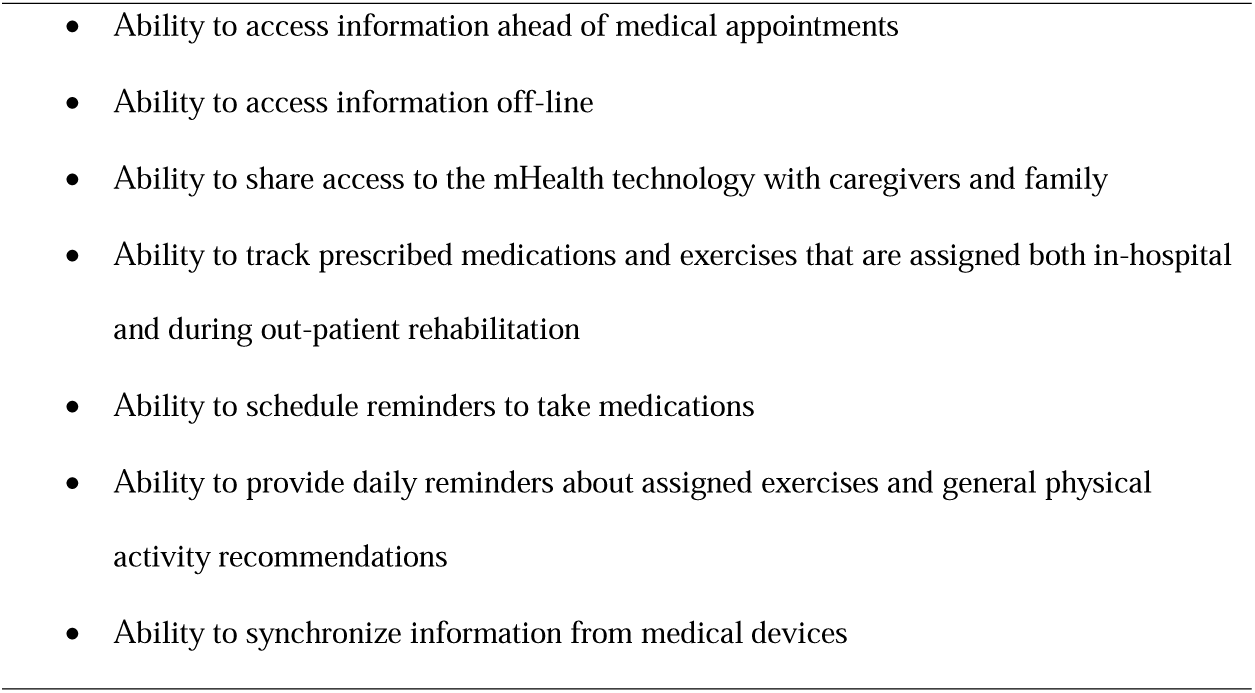
Key messages about mobile health (mHealth) technology design Key messages

### Key messages about mHealth technology content

During discussions of the study’s two open-ended questions and of the generic version of the mHealth technology, content-specific messages related to inclusion of key cardiac surgery team members’ contact information and descriptions of roles, key terms, educational videos, information specific to cardiac surgery procedure, and about travel before/after surgery. When discussing the study’s pre-defined categories of information, key content-specific messages about (a) nutrition related to what to eat, (b) medications included drug interactions, (c) resources included medical devices, and (d) physical activity related to addressing fears, as well as providing information, recommendations, and instructions, were generated by the advisory panel. Specific key messages about mHealth technology content are in Table 3.

**Table 3.**
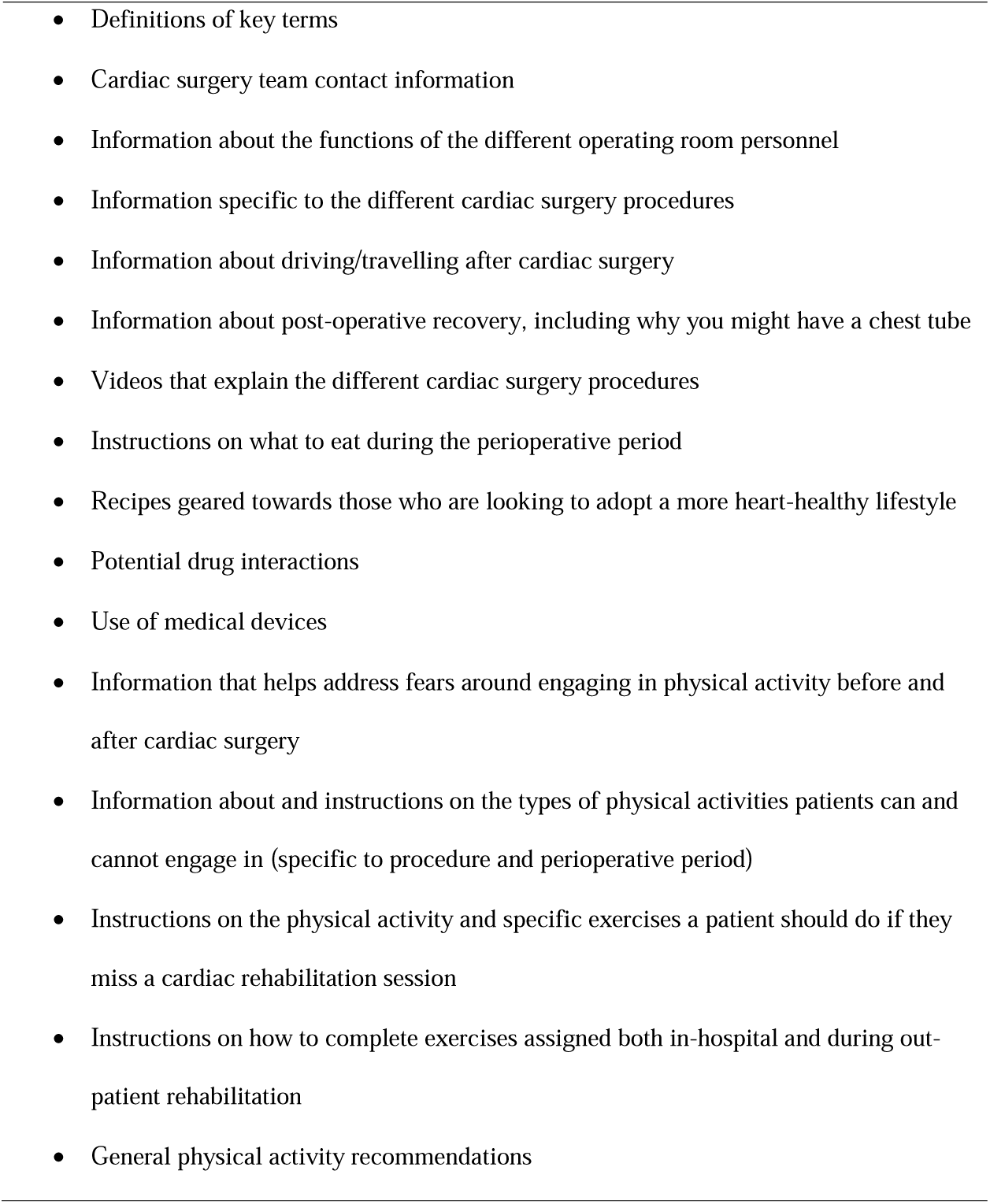
Key messages about mobile health technology content Key messages

### Impacts on mHealth technology design and content

Key messages about the design and content of the mHealth technology were compiled and sent to the mHealth technology developers by the study co-ordinator (DEK). The mHealth technology developers incorporated key messages into the design and content of the mHealth technology, as long as they could be supported by the existing functionalities of the underlying platform. For example, the platform did not support the scheduling of reminders by users, identifying drug interactions, or synchronization with other devices. Verbal and written feedback from the technology developers indicated that the key messages were a richer source of information and provided more guidance than typically received from past clients. In particular, the technology developers noted that key messages resulted in: the integration of a vast range and volume of information and resources, instead of ones primarily focused on surgical information; content geared towards expectations management; and an expanded focus of the mHealth technology to include caregivers and other family, so that these stakeholders may be directly included in the provision of information, allowing them to be better informed, prepare along with the patient, and be involved in recovery planning.

## DISCUSSION

Our findings demonstrate that engaging patients and caregivers in research, through the formation of an advisory panel, yields a rich source of information to guide the design and content of a mHealth technology developed for cardiac surgery patients. These findings are novel since, despite the increased recognition of the importance of involving patients in research, patient engagement remains underutilized in many health research areas, including mHealth technology design [9] and cardiac surgery. Further, while patient input is more regularly sought in the commercial technology arena, it is often obtained through focus groups and/or pilot testing aimed at gathering proprietary data; it is rare that patients and caregivers are engaged as partners and co-creators of the technology.

Several characteristics of the patient engagement activities likely contributed to the gathering of useful information. First, the deliberate intention to create an environment that supported the integration of patients into research, through activities that targeted the guiding principles that underlie patient engagement [10] and as led by a skilled facilitator. Second, a mixture of broad and focused open-ended questions was used, in order to gather spontaneous feedback, as well as feedback related to categories of information based on our previous work. Interestingly, during discussions of the broad, open-ended questions, topics raised tended to concern the potential benefits of the mHealth technology. For example, some of the topics raised by the panel included the technology’s potential to change how patients and caregivers interact with information to better support patient engagement with their health care plan (e.g., through the ability to access information ahead of an appointment to prepare questions or know what to expect, by allowing them to fact-check what they thought they heard during appointments without having to rely on outside sources like internet searches) and the potential for caregivers to become more involved in the patient’s journey. Discussions of the more focused questions produced key messages more directly related to the design and content of the mHealth technology. Third, advisory panel members were selected based on undergoing cardiac surgery within the past two years, ensuring accurate recall of their experience and elaboration on the information that they did and did not receive as part of their patient-provider interaction. This would have had a positive impact on their abilities to contribute to conversations. Further, the advisory panel met on multiple instances, which gave advisory panel members the opportunity to reflect on the study questions and their experiences alone or with caregivers and other individuals that supported them during their patient journeys, and then to bring these reflections back to enrich discussions in subsequent meetings. Finally, the advisory panel included both patients and their caregivers, which provided a breadth of experiences, and turned out to be timely given patients’ statements around the potential of the mHealth technology to allow caregivers to be more involved in the patient’s journey.

With the increase of older adults being offered cardiac surgery, there is an urgent need to provide a high-level of patient-centered value and quality in our perioperative management. The use of evidence-based ERPs has resulted in more rapid and optimal recovery than with traditional perioperative methods (i.e. improved survivorship) in cardiac surgery patients.[22] While published guidelines provide an important framework from which to develop clinical pathways,[23] implementation remains challenging and therefore the protocols are underutilized. It is anticipated that the approach of involving patents and caregivers in the development stage will enable the healthcare team to focus on patient-caregiver value in the development and subsequent implementation phase that will ideally translate to a sustainable process. To this end, the findings from the current work have provided a deeper understanding of patient and caregiver needs pertaining to information delivery about various aspects of peri-operative care and the potential role of mHealth platforms in supporting these recommendations. The impact of this mHealth technology on knowledge delivery and patient adherence with ERPs during the perioperative period of cardiac surgery will be explored in future work.

### Limitations

This study has some limitations to mention. Logistical constraints shaped our patient engagement approach. For example, while we engaged patients and caregivers at specific time points within the study, we did not continually involve them throughout the project as full research co-investigators. Had there been continual engagement, there would have been other points of input and the nature of advisory panel members’ relations with the study would have been different (see Supplementary Material 1). That said, it is important to note that advisory panel members were invited to be co-authors on this manuscript, both to further support the establishment of authentic research partnerships and to ensure that the manuscript accurately reflects their voices and ideas. We also plan to engage advisory panel members further in the re-evaluation and revision of the mHealth technology prior to its adoption as a standard of care tool to be used within the Cardiac Sciences Program at St. Boniface Hospital.

### Conclusions

Patient engagement facilitated the development of a mHealth technology whose design and content were driven by the lived experiences of cardiac surgery patients and caregivers. The result was a detail-oriented and patient-centered mHealth technology that helps to empower and inform patients and their caregivers across the perioperative period of cardiac surgery. Consequently, this mHealth technology has the potential to enhance the delivery and patient experience with ERPs, and ultimately to support better health outcomes.

## Data Availability

No data available as advisory panel members were not made aware beforehand that recordings of meeting proceedings would be publicly available.

## ACKNOWLEDGEMENTS

We acknowledge the contributions of Drs. Arora and Schultz as co-senior authors on this manuscript, as well as of Drs. Chudyk and Schultz as bringing expertise in patient engagement approaches to the larger research project and this manuscript. We would like to thank the members of the Healthy Heart Patient Researchers group for their partnership in this research endeavor, which include Carole Dobson, Carole Hyra, Wayne Dobson and seven other members. Further information about this study, and our other work in the area of patient engagement in research, can be found at www.patientengagementinresearch.ca.

## COMPETING INTERESTS

Mses. Dave, Hyra and Ragheb; Mr. Kent; and Drs. Chudyk, Duhamel, and Schultz have no competing interests to disclose. Dr. Arora reports grants from Pfizer Canada Inc., as well as other (i.e., honoraria) from Mallinckrodt Pharmaceuticals, Abbott Nutrition, and Edwards Lifesciences, outside the submitted work. None of the Authors have any formal relationship (financial or non-financial) with the industry partner in this study.

## FUNDING

This work was supported by a GFT research grant from the Department of Surgery (University of Manitoba). Funding for the development of the study’s mHealth technology included in-kind contributions from BeeWell Health ©. The Authors had full control over the data and the company has not seen a version of this publication prior to submission. Dr. Chudyk’s postdoctoral fellowship was supported by the Department of Family Medicine (University of Manitoba).

## CC BY

No.

## PREPRINTS

This manuscript was deposited as a preprint immediately prior to its submission to Heart.

## ABBREVIATIONS

CABG: Coronary artery bypass grafting
CIHR: Canadian Institutes of Health Research
ERPs: Enhanced recovery protocols
GRIPP2: Guidance for Reporting Involvement of Patients and the Public
mHealth: Mobile health

## Notes

### Clinical Trial

The study was not eligible for registration with a clinical trial registry as it is not a clinical trial.

### Author Declarations

University of Manitoba Research Ethics Board (H2018:458) and the St. Boniface Hospital Research Review Committee Research Ethics Board (RRC/2018/1813).

